# Independent Dentists in the UK have less access to Occupational Health

**DOI:** 10.1101/2020.11.08.20227942

**Authors:** Meena Satish Ranka, Satish Ranka

## Abstract

**Objectives:** Respiratory illnesses like Severe Acute Respiratory Syndrome, Middle East Respiratory Syndrome and the current SARS-CoV-2 virus pandemic are transmitted by respiratory droplets. Certain dental procedures generate aerosols, one of the highest sources of transmission of droplet infections. During the current pandemic, dentists in the UK were initially restricted in their work and now have guidance from NHS England and the Chief Dental Officer for the full resumption of safe and effective routine dental care to patients. Lack of work, impact on income and working in the continual pandemic situation are likely to cause significant mental stress in dentists. Occupational health (OH) can have a vital role to help such dentists remain in work by advising strategies to cope with stress and offering timely evidence-based interventions and adjustments. The aim was to assess if the dentists in the UK had access to OH and whether access to the OH services helped.

**Methods:** A survey link was sent to 200 dentists in the UK after the peak of the current pandemic.

**Results:** 124 dentists responded to the survey. The response rate was 62%. 59% of the dentists in the survey did not have access to OH services in their workplace. Only 15% of dentists working in the independent sector had access to OH services compared to 78% working in the NHS or having NHS contracts. None of the dentists in the survey accessed OH services.

**Conclusions:** Access to OH services for the dentists needs improvement, particularly in the Independent sector in the UK.

**Article Summary:** Strengths and limitations of the study:

- Sample representative of the population studied.
- No observer subjectivity
- Precise results
- Inflexible design, no control group and lack of random sampling

## INTRODUCTION

In the United Kingdom, medical, dental, and healthcare services are provided by government funded organisation called the National Health Services (NHS). Dental care in the UK is provided in NHS hospitals, in practices with an NHS contract and in the independent sector. Dental interventions can include procedures which generate aerosols which are highly infectious and can transmit the virus causing the current pandemic [1]. To prevent the risk of transmitting the pandemic virus during the peak of the pandemic, routine dental work was stopped by the government and only emergency treatment or telephone advice was offered. It is likely that the dentists restricted from clinical work, and those working in the pandemic may have undergone significant stress and experienced mental health symptoms.

Research into the mental health of dentists in the UK started approximately 3 decades ago [2]. Survey research by the British Dental Association in 1996 found 11% of dentists experienced some level of distress [3]. In 2019, survey of 2053 UK dentists concluded that dentistry is a stressful profession [4].

Occupational health, as a specialty, helps in managing long term sickness absence, hazards in workplace, health surveillance and managing blood borne viral infections due to body fluid exposures including needle stick injuries amongst its various other services [5]. Research about OH access/intervention to assist dentists cope with stress and mental health is lacking in the UK and other parts of the world. Further insight into the current level of access to OH services for dentists is important to plan strategies for timely intervention to prevent long term psychological ill-health.

## METHODS

A survey link was sent to 200 dentists in 3 social media dentistry groups and personal communications in the UK as power calculations using Cohen effect size suggested a minimum of 117 responses be achieved for an effect size of 0.9 for this study. The social media dentistry group included a wide variety of professionals like hospital consultants, trainees, general dental practitioners, and specialty dentists, working in the NHS or with NHS contracts and/or the independent sector. Ethical approval was sought from the NHS Health Research Authority. The survey was conducted after the peak of the COVID-19 pandemic in the UK. Appendix 1 has the list of the survey questions. Patient Health Questionnaire – (PHQ-4), an ultra-brief validated tool was used to screen for anxiety and depression symptoms [6]. It had 2 screening questions on anxiety (A1 & A2) and depression (D1 & D2) each. Closed questions about OH access, support, and interventions including with their place of work were asked. Place of work was identified as NHS, Independent sector, and both.

It was not appropriate or possible to involve patients or the public in the design, or conduct, or reporting, or dissemination plans of our research. No patient involved.

Pearson’s chi-squared test of independence was used to determine the mean. Odds ratio was calculated to measure the risk differences between the association of NHS and independent sector practitioners.

## RESULTS

124 dentists took part in the study. The response rate was 62%. The power for sample size of 124 was 0.92. One respondent did not complete the survey and was excluded. Only 45% of the dentists were working at the time of the survey. Of these 18% were working in NHS setting, 27% in the independent sector and 55% in both.

Access to OH: Only 41% (95% CI 0.33-0.50) of the dentists surveyed, had access to OH. OH access was available for 78% (95% CI 0.58-0.90) of dentists working in the NHS, 41% (95% CI 0.30-0.53) working in both (independent sector and NHS) and in 15% (95% CI 0.07-0.32) working solely in the independent sector. With regards to PHQ4 questions related to anxiety and depression screening, 67% (95% CI 0.07-0.32) of anxious dentists, 59% (95% CI 0.45-0.71) of worried dentists, 47% (95% CI 0.34-0.60) of dentists with little interest in doing things and 45% (95% CI 0.32-0.59) of dentists feeling low, had access to occupational health.

Accessing OH services: Surprisingly, none of the dentists including those who were experiencing mental health symptoms and who had access to OH, reported of accessing/seeking OH intervention to improve their psychological health.

However, an interesting finding was that having access to OH services was associated with a 2-3 times lesser risk of developing mental health symptoms for the dentists. (Table 1).

**Table 1.** Odds of the likelihood of development of MH symptoms in dentists as per the PHQ4.

|  | Odds Ratio | 95% Confidence Interval |  |
| --- | --- | --- | --- |
|  |  | Lower | Upper |
| Access to OH, $n=123$ | | | |
| A1 | 2.04 | 0.89 | 4.65 |
| A2 | 2.06 | 0.95 | 4.46 |
| D1 | 2.68 | 1.27 | 5.67 |
| D2 | 2.90 | 1.37 | 6.16 |

\*A1: Anxious, A2: Worried, D1: Little interest, D2: Feeling low

## DISCUSSION

The findings of this survey suggest that a significant number of dentists in this group were not working during the peak of the pandemic. This change in delivery and operation of dental services was due to government restrictions to prevent transmission of the pandemic virus. Just under 60% of the dentists in this study did not have OH access, particularly those working in the independent sector. It was not possible to determine if OH intervention would help as no dentist accessed OH advice. However, the significant finding from this study was that those dentists who had access to OH services were 2-3 times less likely to experience psychological symptoms.

A recent survey by Toon et.al of 1513 dentists in the UK found statistically significant relationship (p<0.01, β=0.67) between work related stress and burnout in UK dentists amongst other factors [7]. Research in other parts of the world have highlighted similar findings over the last two decades [8,9]. OH can provide timely intervention for fitness to continue working by suggesting restrictions/adjustments, referral to employee assistance programme to access counselling services and ongoing review [10]. In the UK, dentists working in an NHS setting usually have an inhouse OH service provision compared to the independent sector where the access to the OH is usually a paid service. There may be a lack of awareness of the various services offered by OH, the associated costs and ease of access may be some of the factors contributing to the differences in access to OH services between independent sector and the NHS. This topic needs further investigation.

Limitations of this survey include a snapshot view of dentists in the UK at the time of the pandemic. Respondents may have an exacerbated emotional response due to the mortality and morbidity associated with severe Covid 19 disease, loss of earnings, lack of a routine, hypervigilance, social distancing in the pandemic situation; all of these can lead to a response bias. However, access to OH is a factual question, unlikely to be biased by an external factor. As none of the dentists accessed the OH services, precise estimate of risk reduction in the mental health symptoms in dentists who had OH access remain a topic of future research.

In conclusion, findings of this study suggest that there is a need for improvement in access to the OH services for dentists, particularly in the independent sector. We support the idea of improving OH access for the dentists in workplace to receive timely and evidence-based interventions in line with the current Society of Occupational Medicine, UK campaign of Universal OH access.

## Data Availability

Data will be available on request

## Contributorship Statement

Both the authors were involved in the design, execution, data collection, analysis and writing of the manuscript.

## Competing interests

Nil

No funding was needed for the study.

Data will be shared on request.

### Appendix 1: Survey Questions and responses

1. Over the last 2 weeks, how often have you been bothered by feeling nervous, anxious or on edge? Not at all/Several days/more than half the days/ nearly every day. (A1)
2. Over the last 2 weeks, how often have you been bothered by not being able to stop or control worrying? Not at all/Several days/more than half the days/ nearly every day. (A2)
3. Over the last 2 weeks, how often have you been bothered by little interest or pleasure in doing things? Not at all/Several days/more than half the days/ nearly every day. (D1)
4. Over the last 2 weeks, how often have you been bothered by feeling down, depressed or hopeless? Not at all/Several days/more than half the days/ nearly every day. (D2)
5. Do you have access to Occupational health? Yes/No/Other.
6. Have you accessed Occupational health for psychological wellbeing in the current pandemic? Yes/No/Not applicable
7. Did Occupational Health intervention help you with your psychological wellbeing? Yes/No/Not applicable
8. Are you currently working? Either in dentistry or redeployed? Yes/No
9. You practice: Exclusively in NHS/Exclusively in independent sector/Both in NHS and independent sector/Other.

